# Host serine proteases and antiviral innate immunity as potential therapeutic targets in influenza A virus infection-induced COPD exacerbations

**DOI:** 10.1101/2025.02.02.25321231

**Authors:** Haiqing Bai, Melissa Rodas, Longlong Si, Yuncheng Man, Jie Ji, Roberto Plebani, Johnathan D. Mercer, Rani K. Powers, Chaitra Belgur, Amanda Jiang, Sean R. R. Hall, Rachelle Prantil-Baun, Donald E. Ingber

## Abstract

Lung manifestations of chronic obstructive pulmonary disease (COPD) are often exacerbated by influenza A virus infections; however, the underlying mechanisms remain largely unknown and hence, therapeutic options are limited. Using a physiologically relevant human lung airway-on-a-chip (Airway Chip) microfluidic culture model lined with human airway epithelium from COPD or healthy donors interfaced with pulmonary microvascular endothelium, we observed that Airway Chips lined with COPD epithelium exhibit an increased sensitivity to influenza virus infection, as is observed clinically in COPD patients. Differentiated COPD airway epithelial cells display increased inflammatory cytokine production, barrier function loss, and mucus accumulation upon virus infection. Transcriptomic analysis revealed gene expression profiles characterized by upregulation of serine proteases that may facilitate viral entry and downregulation of interferon-related genes associated with antiviral immune responses. Importantly, treatment of influenza virus-infected COPD epithelium with a protease inhibitor, nafamostat, ameliorated the disease phenotype as evidenced by dampened viral replication, reduced mucus accumulation, and improved tissue barrier integrity. These findings suggest that targeting host serine proteases may represent a promising therapeutic avenue against influenza-afflicted COPD exacerbations.

## INTRODUCTION

Chronic obstructive pulmonary disease (COPD) is a common disease with high global morbidity and mortality, which affects one in every ten individuals worldwide (*1*). COPD is characterized by an enhanced inflammatory response of the lung airway epithelium to inhaled gases and particles, particularly cigarette smoke, which leads to gradual airflow limitation and decline of lung function. However, fewer than 50% of smokers develop COPD, suggesting that host factors also may contribute to disease pathogenesis (*2*). Patients with COPD usually experience long-term progressive damage to their lungs but also frequently experience acute episodes of worsening in disease symptoms, known as exacerbations. Respiratory viral infections, such as those caused by influenza viruses and SARS-CoV-2, are common triggers for exacerbation and contribute significantly to hospitalizations (*3*).

Despite such high prevalence, the underlying mechanisms that contribute to the enhanced susceptibility to viral infection in COPD patients remain poorly elucidated. Respiratory viruses, such as influenza A viruses (IAVs), preferentially target airway epithelial cells, leading to epithelial cell sloughing, microvascular dilatation, lung edema, immune cell recruitment, and associated inflammatory responses, which further impairs lung function in COPD patients. Due to this hyper-reactive response to infection, common antiviral drugs that are effective in healthy patients, such as oseltamivir, are not clinically useful in management of influenza in COPD (*1*). Annual vaccinations are currently the only recommended strategy for care of COPD patients, but these recommendations are based largely on observational studies (i.e., not randomized controlled trials), and the efficacy of influenza vaccines is relatively low due to rapid mutation of the viral genome. Thus, there is an urgent need to better understand the molecular basis of COPD exacerbations by viral infection as well as for new therapeutics that can mitigate infection and attenuate acute deterioration of lung function in these patients.

Here, we describe how we found that a human lung airway-on-a-chip (Airway Chip) microfluidic culture model that has been previously shown to faithfully recapitulate lung pathophysiology (*4*) also replicates the enhanced sensitivity of the COPD lung airway to IAV infection compared to healthy lung. We extended these findings to more donors using higher throughput Transwell cultures and analyzed viral load, cytokine secretion, and transcriptomic profiles in airway epithelial cells to explore potential mechanisms that result in increased susceptibility to influenza infection in COPD patients. These studies revealed an increase in the expression of genes that encode serine proteases which activate viral entry and a decrease in interferon (IFN)-related genes involved in host antiviral innate immunity. Moreover, we demonstrate that virus-induced COPD exacerbations can be suppressed in vitro by using a protease inhibitor drug that targets the host rather than the virus.

## RESULTS

### Human COPD Lung Airway Chips are more sensitive to influenza virus infection

COPD patients are known to be particularly susceptible to respiratory viral infections, however, whether this is due to a direct sensitivity of the airway epithelium or an abnormal immune or inflammatory response remains unknown. To directly address this question, we leveraged a human lung Airway Chip that has been previously shown to replicate many human host responses to virus infection in vitro (*5*). The Airway Chip is an optically clear silicone rubber device that contains two parallel microchannels separated by an extracellular matrix (ECM)-coated porous membrane lined on one side by primary human lung airway epithelial cells cultured under an air-liquid interface (ALI), and on the other side by human pulmonary microvascular endothelial cells (HPMVECs) grown in the presence of continuous medium flow to mimic vascular perfusion (**Fig. 1A**). This 3D multi-tissue model faithfully reproduces many structural and functional features of the human upper airway, including differentiation of all cell lineages in normal proportions, mucociliary movement, and tissue barrier integrity, and thus allows investigation of various facets of viral infection of the upper airway, including viral tropism, host cytokine production, immune cell responses, drug efficacy, and viral evolution of drug resistance (*4, 6*). When populated with lung cells from diseased patients, the Airway Chip also enables faithful disease modeling, as recently demonstrated for cystic fibrosis (*7*).

**Fig. 1.**
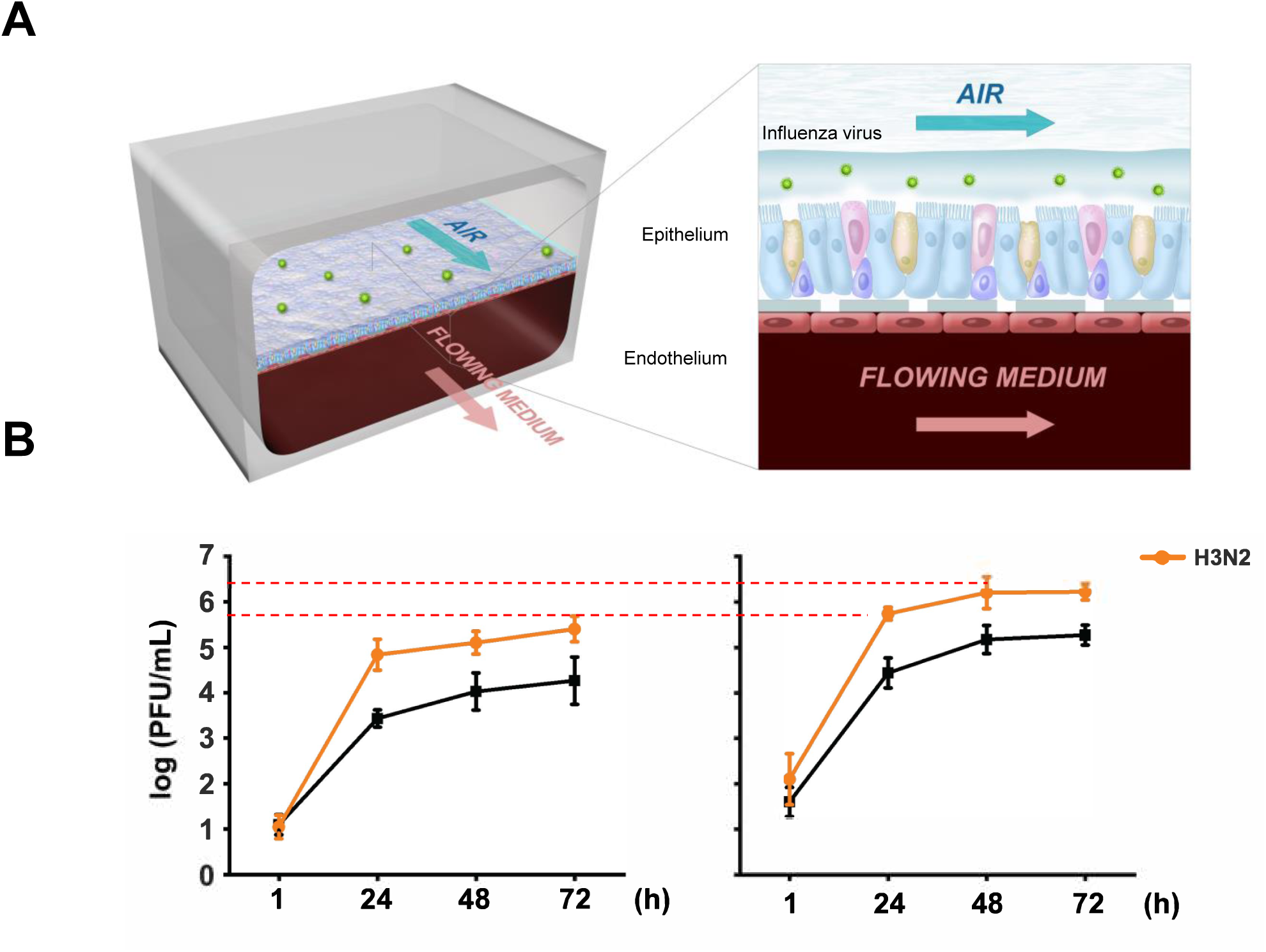
Increased influenza virus replication in COPD Airway Chip. **(A)** Schematic illustration of a human airway chip. The human airway chip is a two-channel microfluidic device with an air channel and a blood channel. Human primary airway cells cultured at an air-liquid interface line the air channel; human pulmonary endothelial cells line the blood channel. To replicate the airborne route of transmission, influenza virus is introduced into the air channel. (**B**) Viral titers for Influenza A/WSN/33 (H1N1) and influenza A/Hong Kong/8/68/ (H3N2) virus replication in a 3-day time course in the Human Lung Airway Chip. N=3 independent Chips from 2 donors for each group. Both strains were inoculated at MOI = 0.01.

Here, we adopted a similar strategy to examine the direct responses of healthy versus COPD lung airway epithelium to IAV infection. When inoculated with influenza A/WSN/33 (H1N1) or HK/68 (H3N2) strains of IAV, the COPD Airway Chip exhibited more rapid viral replication and peak viral loads that were more than 10-fold higher than those observed in healthy chips (**Fig. 1B)**. This is consistent with the clinical observation that patients with COPD often experience longer days of sickness and more severe symptoms (*8*).

### Increased influenza virus replication and inflammation in COPD airway epithelium

To carry out similar studies in a higher throughput manner in order to extend our findings to more donors, we cultured human airway basal stem cells isolated from 4 healthy donors and 4 COPD donors (**Table 1**) under air-liquid-interface (ALI) on the top surface of the porous membrane in 24-well Transwell inserts (*9*) for 3 weeks. Under these conditions, the epithelium generates a pseudostratified layer of cells that include all major cell types in the human airway, including basal cells, ciliated cells, goblet cells, and club cells. We also seeded human microvascular endothelial cells on the bottom surface of the same membrane 2 days before infection with IAV/WSN/33 (H1N1) virus for 18 hours. Similar to our results with the Airway Chip, these studies suggested that H1N1 infection generates a higher viral load in COPD airway epithelium compared to healthy tissue, as determined by qPCR analysis; however, there was greater variability in this study involving 4 different donors and thus, the results did not reach statistical significance (**Fig. 2A**). The variation observed between donors highlights the heterogeneity of COPD but also may reflect different disease stages of these patients.

**Fig. 2.**
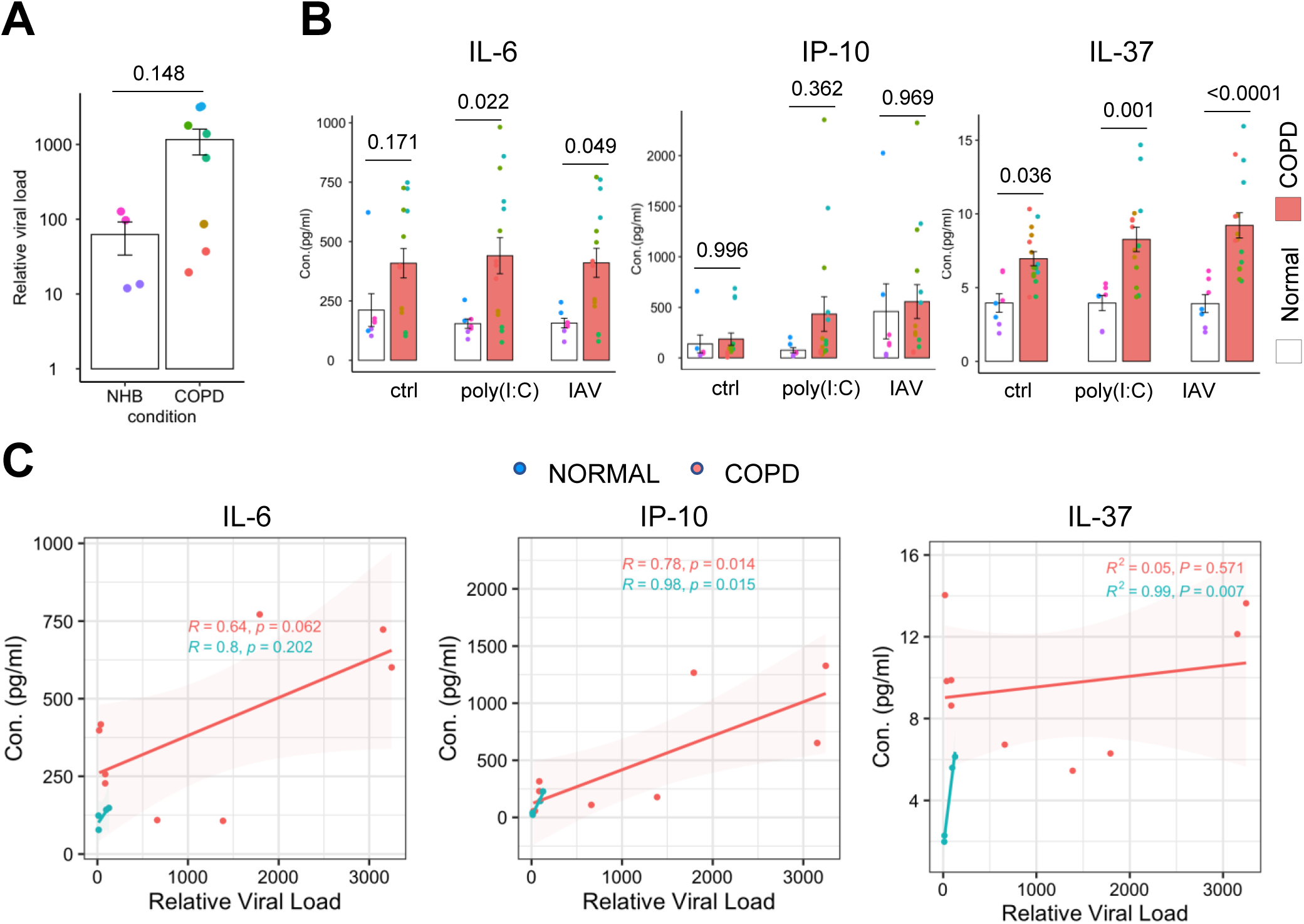
Increased viral infection and cytokine production in the COPD airway epithelium. **(A)** Relative influenza viral load from the apical washes of the Transwell at 18 hours post infection. Mann-Whitney test. (**B**) Protein levels of IL-6, IP-10, and IL-37 measured in the basal medium. Each color in the dot plot represents a different donor and same color represents same donor but different biological replicates. ANOVA and Šidák multiple comparisons test. *p<0.05, **p<0.01, ***p<0.001, ****p<0.0001. (**C**) Pearson correlations between relative viral load and cytokine concentrations.

**Table 1.**
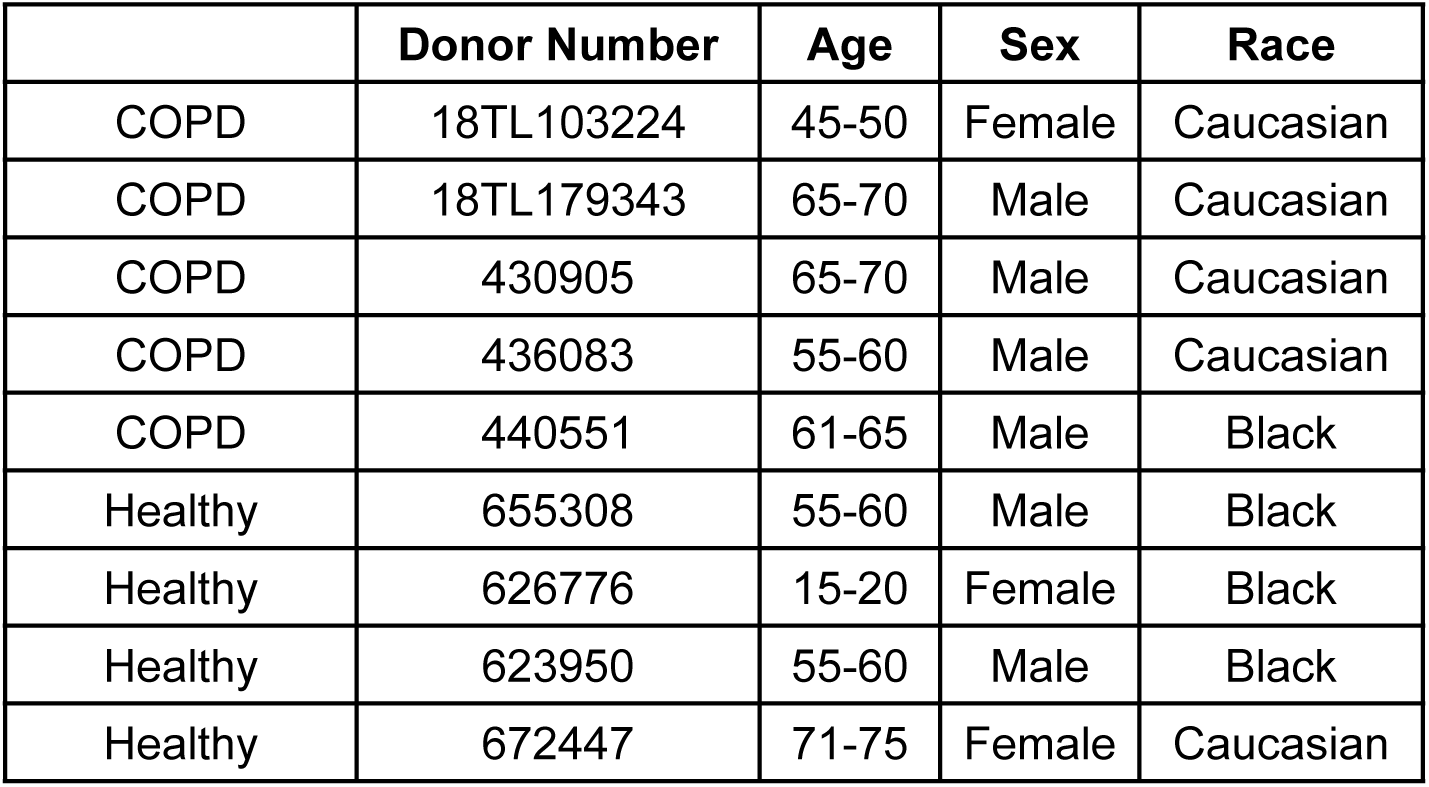
Demographics of airway basal cells from healthy donors or COPD patients.

COPD is characterized by chronic airway inflammation, which is accompanied by increased secretion of pro- and anti-inflammatory cytokines and chemokines. Consistent with this, we detected significantly (P < 0.001) higher levels of IL-6 and IL-37 under basal conditions in the COPD cultures, as well as following infection with influenza H1N1 or even when treated with the pro-inflammatory viral double-stranded RNA analog poly(I:C) (**Fig. 2B**) whereas no significant differences were observed for IL-8, RANTES, or MCP-1(**Fig. S1**). While the difference between average IP-10 values were not statistically significant due to high variability in levels between donors (**Fig. 2B**). Pearson correlation analysis between viral load and cytokine concentrations revealed significant positive correlations for IP-10 as well as IL-6 (**Fig. 2C**), suggesting that these cytokines may contribute to the hyperinflammatory state observed in acute exacerbation of COPD during IAV infection. The level of IL-37 correlates with viral load in healthy controls but not in COPD. Taken together, these findings demonstrate that COPD airway epithelium is hyper-reactive to IAV infection in terms of inflammatory cytokine production and that this correlates with the increased viral load that is observed in these cells compared to healthy controls.

### COPD airway epithelium has an intrinsic enhanced susceptibility to influenza virus entry

We next explored the potential mechanism of increased viral infection in COPD airway epithelium. Successful entry of IAV into host cells requires the cleavage of influenza hemagglutinin (HA) by host proteases that are expressed in the human airway, including transmembrane serine protease 2 (TMPRSS2) and transmembrane serine protease 4 (TMPRSS4) (*10*). In contrast, plasminogen activator inhibitor 1 (SERPINE1) antagonizes IAV virus maturation by inhibiting the activity of these proteases (*11*). We found that the COPD epithelial cells express significantly higher levels of TMPRSS4 and lower expression of SERPINE1 compared to healthy epithelium (**Fig. 3A)**. We did not detect any difference in expression of TMPRSS2 although cathepsin B (CTSB) expression was also higher in the COPD cells (**Fig. S2**). Therefore, COPD airway epithelium may have an intrinsic enhanced susceptibility to IAV entry.

**Fig. 3.**
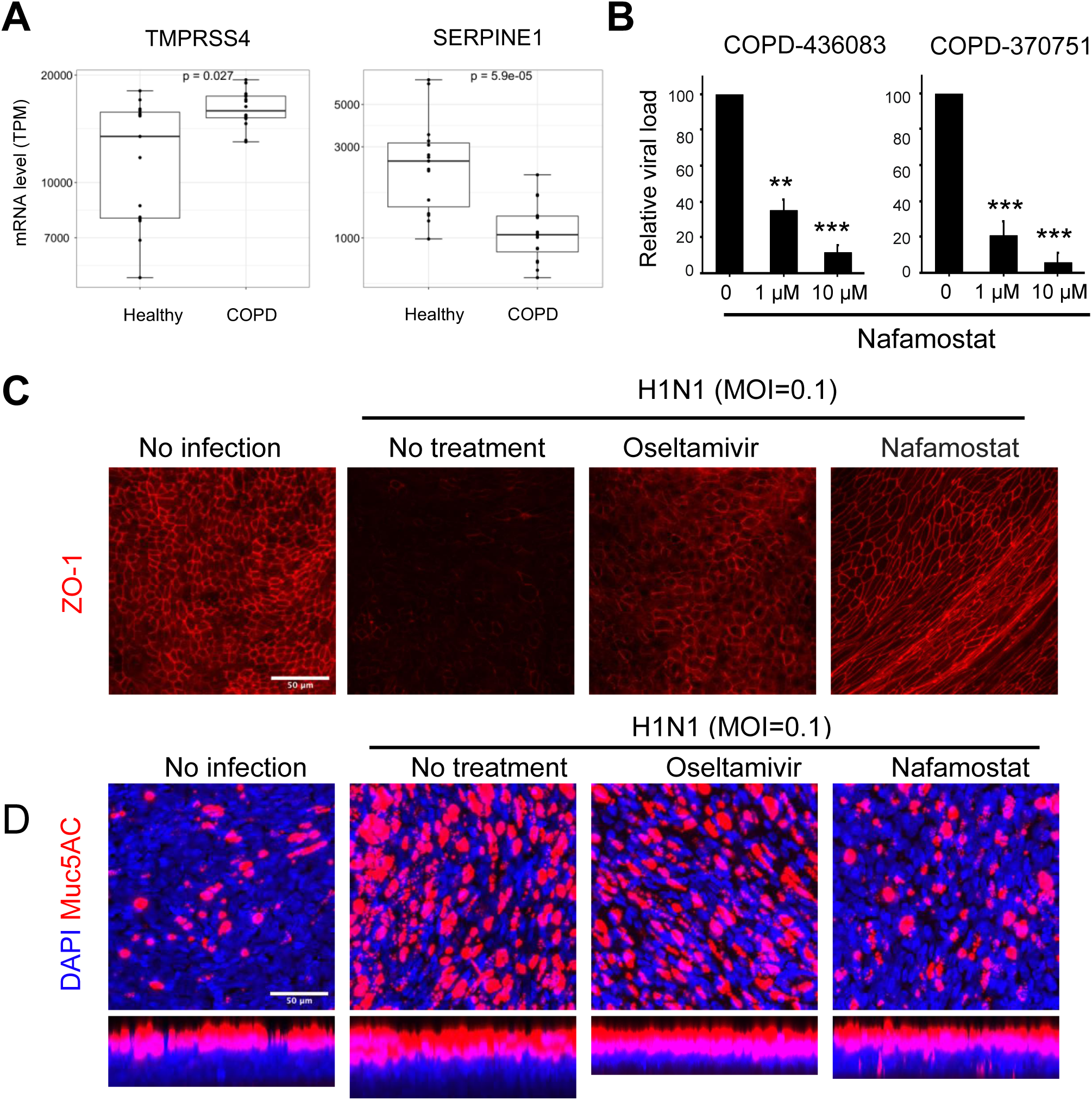
Nafamostat as a potential treatment for influenza infection in COPD. (A) mRNA levels of influenza-activating host protease TMPRSS4 and anti-protease SERPINE1. Mann-Whitney test. (**B**) Relative viral load in apical washes of cells treated with 1 µM or 10 µM Nafamostat at 24 hours before infection with H1N1 influenza virus at MOI = 0.01. One-way ANOVA with Tukey test for multiple comparison, **p<0.01, ***p<0.001. (**C and D**) Immunofluorescence micrographs showing preservation of ZO1-containing tight junctions (**C**) or attenuation of influenza-induced mucus production (**D**) in COPD airway epithelium infected with H1N1 (MOI = 0.01) at 48 h post infection by treatment with nafamostat (1 µM) or oseltamivir (OSV) (1 µM). scale bar, 50 µm.

### Host serine protease inhibition suppresses virus infection and mucus production

We have previously demonstrated that the serine protease inhibitor and an anticoagulant drug, nafamostat, can inhibit IAV infection in healthy Lung Airway Chips, and that it can double the therapeutic time window of the antiviral drug oseltamivir when given in combination (*4*). Given that therapeutic strategies for COPD exacerbation by IAV infection are limited, we explored whether nafamostat also can suppress viral infection and tissue injury in COPD epithelium. We found that nafamostat significantly reduces viral loads when administered 24 hours prior to infection at both 1 µM and 10 µM, and similar results were obtained with cells from two different COPD donors (**Fig. 3B)**. Moreover, treatment with nafamostat at 1 µM also prevented the epithelial barrier disruption (removal of epithelial ZO-1-containing tight junctions) that occurs following virus infection, and this protective effect of inhibiting host proteases was similar to that induced by treatment with the direct acting antiviral drug oseltamivir, as indicated by immunofluorescence microscopy (**Fig. 3C)**. Influenza virus infection also results in increased production of airway mucus in the COPD epithelial cells, and this was totally prevented by nafamostat as well, but interestingly, not by oseltamivir (**Fig. 3D)**. Thus, our results demonstrate that an imbalanced expression of host proteases and antiproteases may contribute to increased influenza virus infection and associated structural changes in the COPD airway, and that nafamostat or other serine protease inhibitors might offer a potential treatment for influenza virus infection-induced exacerbations in COPD patients.

### Attenuated antiviral innate immunity in COPD epithelium

As changes in proteases are only one component of the host defense response to infection, we performed RNA-seq analysis of the differentiated healthy versus COPD airway epithelium grown in Transwell inserts without viral infection (**Fig. 4A**). Examination of cell marker genes showed that COPD airway epithelium expressed higher levels of the basal cell marker keratin 5 (KRT5)(**Fig. 4B**), consistent with previous findings demonstrating exhaustion of airway basal progenitor cells (*12*) and increased numbers of basal cells epigenetically committed to form distinct metaplastic lesions in lungs of COPD patients (*13*). In contrast to previous results, we found increased expression of biomarker genes for club cells (Secretoglobin Family 1A Member 1, CC10; SCGB1), ciliated cells (Forkhead Box J1; FOXJ1), and pulmonary neuroendocrine cells (tubulin beta 3 class III; TUBB3) in COPD airway epithelium while no differences were observed for the mucus cell marker MUC5B at the mRNA level (**Fig. 4B**). A volcano plot (**Fig. 4C**) revealed that there are more genes downregulated than genes upregulated (1703 vs 134) in COPD epithelium compared to healthy, which corroborates recent evidence indicating that COPD is associated with DNA hyper-methylation and epigenetic silencing of transcription (*14*).

**Fig. 4.**
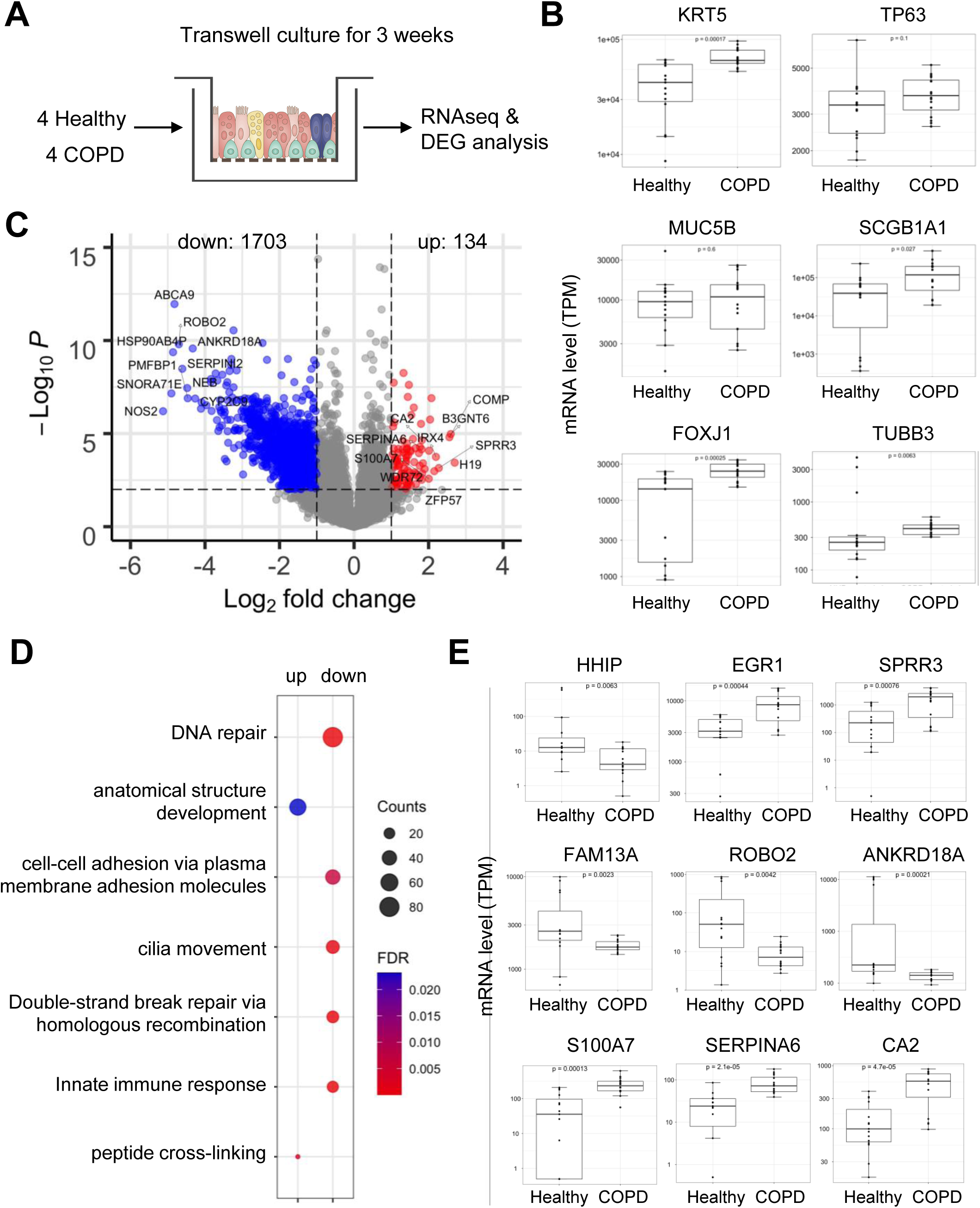
RNA-seq analysis identifies differentially expressed genes in COPD under sterile condition. (**A**) Diagram illustrating reconstituted human bronchial airway epithelium for RNA-seq analysis. (**B**) Box plots showing mRNA levels of major cell marker genes in the airway epithelium. Wilcoxon signed-rank test. TPM, transcripts per million. (**C**) Volcano plots showing differentially expressed genes in COPD vs healthy; P value threshold = 0.01; fold change threshold =2. (**D**) Gene Ontology (GO) enrichment of biological pathways that are upregulated (up) or downregulated (down) in COPD. (**E**) Box plots showing mRNA expression of genes previously shown to be involved in COPD from RNA-seq or GWAS study or (B) new genes from this study. Wilcoxon signed-rank test.

Gene Ontology (GO) enrichment analysis revealed that upregulated genes belong to anatomical structure development and peptide cross-linking, while downregulated genes belong to pathways previously known to be dysregulated in COPD (*8*), including DNA repair, cell adhesion, cilia movement, and innate immune response (**Fig. 4D** and **Table S1**). Finally, we examined the expression of several known genes found to be associated with COPD based on GWAS studies (*15*), including HHIP (Hedgehog Interacting Protein), FAM13A (Family With Sequence Similarity 13 Member A), ROBO2 (Roundabout Guidance Receptor 2), and ANKRD18A (Ankyrin Repeat Domain 18A). We found these genes to be significantly decreased in the COPD airway epithelium (**Fig. 4E**), while other genes including EGR1 (Early Growth Response 1) and SPRR3 (Small Proline Rich Protein 3) were increased. In addition to these known marker genes, we also observed several previously unreported genes that were upregulated, including S100A7 (S100 Calcium Binding Protein A7), SERPINA6 (Serpin Family A Member 6), and CA2 (Carbonic Anhydrase 2) (**Fig. 4E**), which may serve as potential molecular markers for COPD. These results demonstrate that isolated airway basal cells from both healthy donors and COPD patients can differentiate into mature cell types and the reconstituted diseased epithelium exhibits dysregulation of biological pathways specific to the COPD etiology.

We next expanded the RNA-seq analysis to conditions when cells were infected with influenza H1N1 virus or treated with poly(I:C). Principle component analysis (PCA) revealed that donor differences contribute to the biggest sample variances (**Fig. S2**), consistent with the heterogenous responses to infection we observed (**Fig. 2A**). Differential gene expression analysis showed that H1N1 virus infection resulted in significant upregulation of many genes in both groups (**Fig. 5A,B**), but the expression of many more genes is enhanced in the healthy cells compared to the COPD group (90 vs. 52). GO enrichment analysis showed that most of these genes belong to antiviral defense and type I/III interferon (IFN) responses (e.g., STAT1, ISG15, DDX58, MX1) (**Fig. 5C**). This response to viral infection makes sense given that innate immune responses mediated by type I and III IFNs constitute the first-line of host protection against various pathogens including IAV (*16*).

**Fig. 5.**
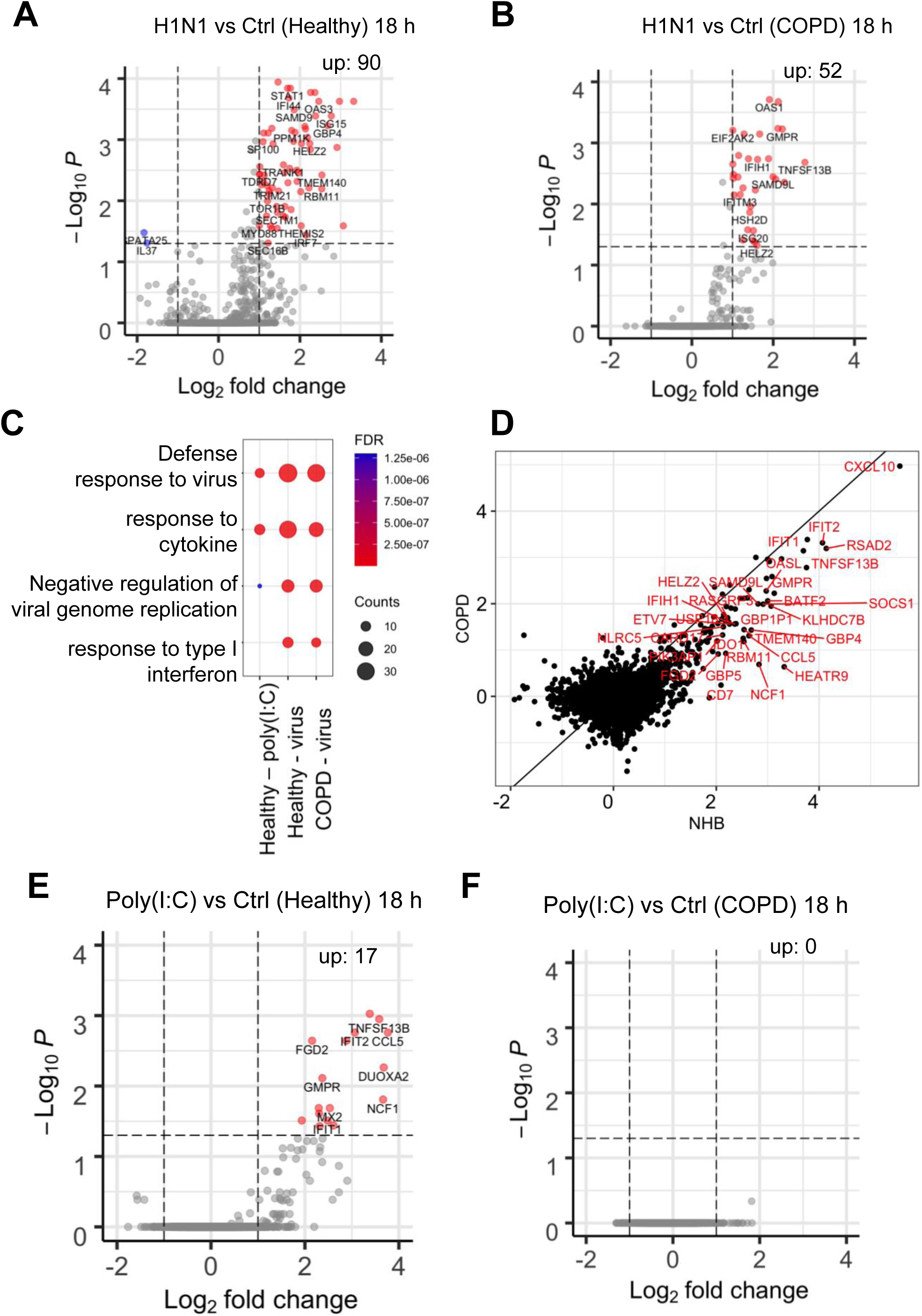
RNA-seq analysis identifies differential host responses to influenza infection in COPD. (**A**) Volcano plot showing DEGs in H1N1 infected (MOI=0.1) group vs control in the healthy donors. (**B**) Volcano plot showing DEGs in H1N1 infected (MOI=0.1) group vs control in the COPD donors. (**C**) Dot plots showing gene ontology enrichment of biological pathways that are upregulated genes in **A** and **B**. (**D**) Scatter plot showing gene expression fold changes of ISGs by influenza infection in COPD vs NHB. (**E**) Volcano plot showing DEGs in the poly(I:C) treated (10 pmol/mL) group vs control in the healthy donors. (**F**) Volcano plot showing DEGs in the poly(I:C) treated (10 pmol/mL) group vs control in the COPD donors.

These findings raised the possibility that the observed hypersensitivity to IAV infection in COPD airway epithelium could be in part due to an impairment of this antiviral IFN I/III response. Indeed, when we compared the expression of these genes in response to influenza H1N1 infection in healthy versus COPD epithelium, the IFN-stimulated genes (ISGs) were suppressed in COPD cells compared to healthy airway epithelium (**Fig. 5D**). This is further supported by the finding that treatment with a low concentration (10 pmol/mL) of the viral mimic Poly(I:C) resulted in upregulation of many genes involved in innate immune defense in the healthy epithelium, but not in the COPD cells (**Fig. 5E,F**). Finally, we performed a Pearson correlation analysis of viral titers and mRNA expression of 6 different ISGs. In healthy airway epithelium, IAV infection resulted in robust stimulation of all the ISGs examined, whereas there was a much weaker induction in COPD epithelium (**Fig. 6**). Taken together, these results indicate that the COPD epithelium exhibits an impaired antiviral innate immune response, which may contribute to unchecked replication of IAV as well as increased inflammation, mucus production, and tissue injury in the airway.

**Fig. 6.**
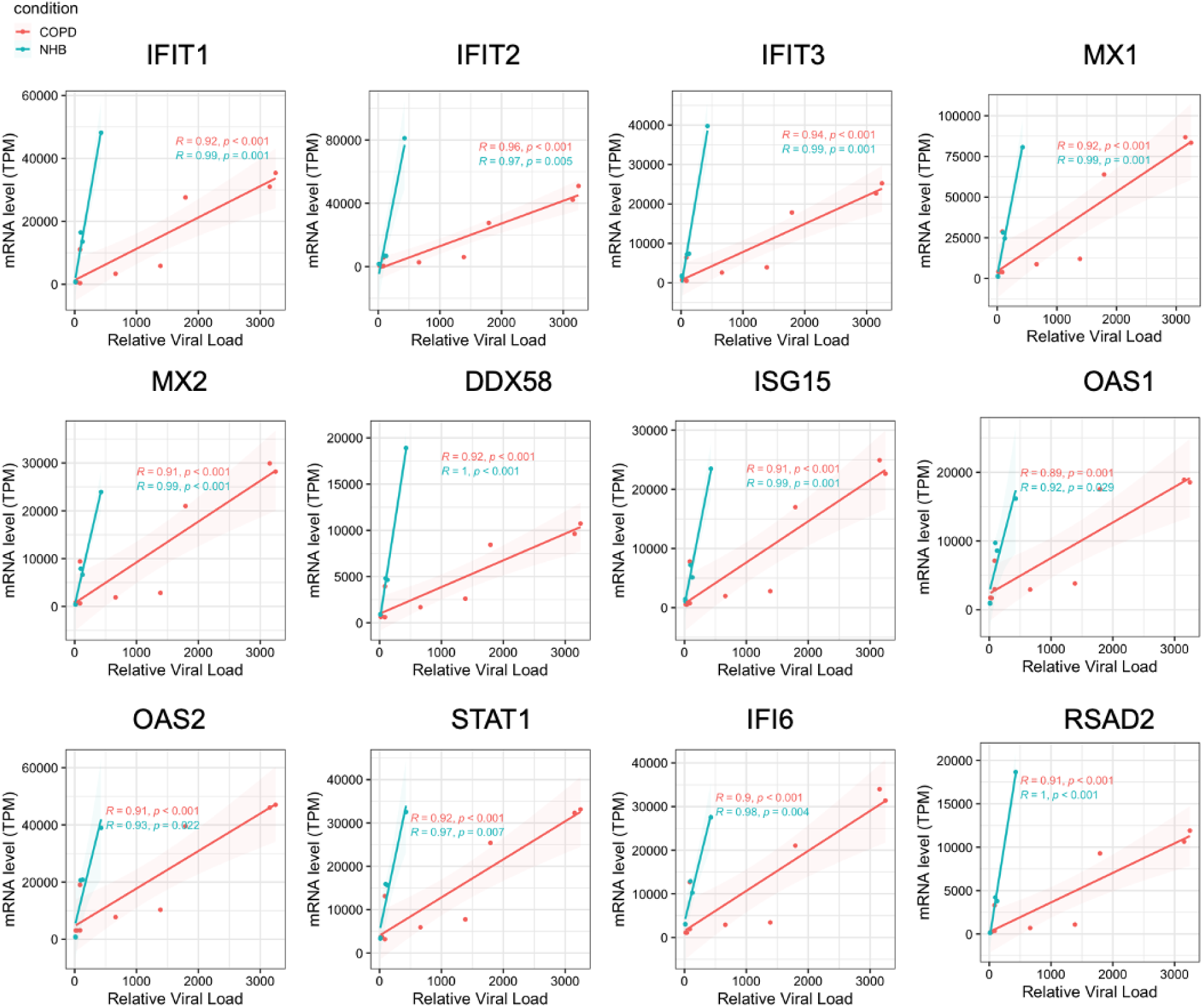
Delayed antiviral defense response in COPD. Pearson correlations between relative viral load and mRNA level of ISGs.

## DISCUSSION

In this study, we sought to gain greater insight into the molecular basis of viral infection-induced exacerbation of COPD in the human lung airway by studying this process in vitro. Using two different organotypic human lung airway culture models (Organ Chip and Transwell), we show that the human COPD epithelium is hyper-susceptible to IAV virus infection and hyper-reactive in terms of inflammation, mucus production, and cell injury responses. These findings are consistent with observations from 2D cultures of primary bronchial epithelial cells infected with H1N1 or H3N2 strain of IAV (*17*), as well as from a rhinovirus Infection model of the human airway (*18*). Deficient innate antiviral immunity has been proposed to be a major contributor to increased susceptibility to viral infection in COPD patients (*1*), and our transcriptomic analyses of epithelium infected with influenza H1N1 virus or treated with pro-inflammatory poly(I:C) support this concept. Moreover, similar findings were found in a recent study in which COPD airway epithelium was infected with rhinovirus-A1 (*18*). Moving forward, it will be important to explore whether the impairment in host innate antiviral immunity results from epigenetic alterations by cigarette smoking, as this has been shown to suppress production of type I IFN and ISGs (*1*). Thus, pulmonary administration of recombinant interferon or interferon-inducing reagents may serve as a viable treatment for influenza infection in COPD patients.

However, our study goes further and suggests that the increased susceptibility of COPD epithelial cells to virus infection also may be due to an imbalance between virus entry enhancing serine proteases (e.g., TMPRSS4, CTSB) and inhibitory antiproteases, such as SERPINE1, which is consistent with our previous results that cigarette smoke in COPD chips upregulates the expression of host proteases TMPRSS11E and TMPRSS11F (*19*). Notably, RNAseq of epithelial brushings and bronchial biopsies from COPD patients also revealed elevations in host proteases, which have been postulated to contribute to enhanced infection with SARS-CoV-2 (*20*). Importantly, based on this observation, we showed that a serine protease inhibitor drug currently used as an anticoagulant, nafamostat, could potentially be repurposed as a treatment for influenza virus infections in COPD patients. We previously showed that nafamostat can synergize with the first-line antiviral drug for influenza, oseltamivir, and that it does not induce drug resistance because it targets the host rather than the virus (*6*). Therefore, nafamostat or other serine protease inhibitors may represent novel treatment options for acute exacerbation of COPD due to viral infection.

Genetic factors are important determinants of COPD in addition to aging, history of smoking, and other environmental hazard exposure. GWAS-based analysis remains the major way to understand COPD genetics and it has led to the identification of many COPD-associated gene loci (*15*). However, the reproducibility of different studies is low and much of the estimated heritability for COPD remains unexplained. On the other hand, transcriptomic profiling using microarrays or RNA-seq has found little overlap among COPD candidate genes identified in different studies, most likely because the cellular composition of tissues analyzed and the diversity of COPD diseases stages are likely the largest drivers of sample variability (*21*).

In this study, we used well-defined in vitro culture systems to regenerate differentiated human airway epithelium using patient-derived primary basal cells. Our results show that COPD basal cells can differentiate into mature cell types (e.g., goblet cells, ciliated cells) in vitro. The expression of the basal cell marker KRT5 is increased in COPD consistent with the recent finding from an in vitro colony-forming assay, which indicates that COPD progenitor cells produce an epithelium with increased basal cells (*12*). However, in contrast to this past study, we did not observe increased goblet cells or decreased ciliated cells under baseline culture condition. This discrepancy may be due to the heterogeneity of donors (such as age, sex, ethnicity) or the differentiation approach we used, as recent evidence from single-cell RNA-seq suggests that human basal cells cultured using the colony-forming assay possess higher colony frequency potential and have altered gene expression compared with primary basal cells (*22*). Additionally, we found that IAV appears to replicate more efficiently in the human Airway Chip and result in higher levels of cytokine production than in the static Transwell model (*23*). This might be due to enhanced epithelial-endothelial crosstalk on the chip, or increased ciliated cell differentiation, as indicated by a recent study (*23*). Future investigation is needed to determine the underlying mechanisms.

Our transcriptional analysis suggests that most genes that are differentially expressed in COPD versus healthy epithelium are downregulated, which is consistent with the results from profiling COPD-associated genes in nasal epithelium (*24*). Most of the downregulated genes relate to pathways that are implicated in COPD pathogenesis, such as DNA repair, cell adhesion, cilia movement, and innate immune response (*25*). Importantly, major COPD-associated genes that were previously identified from a GWAS study also show altered expression in our analysis. These include HHIP (Hedgehog interacting protein), a gene that has been consistently associated with COPD (*15*) and whose haploinsufficiency predisposes mice to develop age-related emphysema (*26*). Our data also indicate that the expression of FAM13A is lower in airway epithelium of COPD patients compared with healthy controls, which contradicts results from a murine model of cigarette smoke-induced COPD (*27*); importantly, however, it is consistent with a recent finding from human patients (*28*). Moreover, SERPINA6 (Serpin Family A Member 6, an antitrypsin), whose variant is associated with COPD, also showed increased expression in our study. This gene could serve as a potential novel marker for COPD and its functional relevance in lung function and COPD pathogenesis warrant further investigation. In addition to increased production of the well-characterized cytokine IL-6, we also detected increased levels of anti-inflammatory IL-37 in COPD epithelium under baseline (non-infected) conditions. Increased expression of this inflammasome inhibitory molecule also has been demonstrated in bronchial alveolar lavage from COPD patients (*29*).

Despite our novel findings, our work has several limitations. First, although we studied multiple donors, the total number (4) is still small. Given the highly heterogeneous nature of COPD, future research is required to include more donors of varying ages, sexes, and ethnic origins. Another limitation is that because we obtained our primary cells from commercial sources, we lack background information for these donors (e.g., smoking history, work environment) including their Global Initiative for Chronic Obstructive Lung Disease (GOLD) grading stage. Finally, we used cells from the bronchial airway, but COPD is a complex lung disease that is associated with abnormal lung injury, repair, and inflammation in other parts of the lung, including the small airway and alveolus as well. Future research will be required to examine whether impaired innate immunity also occurs in these regions. Nonetheless, our study establishes a paradigm for studying COPD exacerbations induced by IAV infection in vitro and shows that host serine proteases and host innate immunity may be a potential targets for therapeutic intervention in patients with COPD exacerbations caused by viral infection.

## Data Availability

All data produced in the present study are available upon reasonable request to the authors

## Acknowledgements.

We thank Zach Herbert at the Molecular Biology Core Facilities at Dana-Farber Cancer Institute for help in RNA sequencing. This work is supported by funding from NIH (1-UG3-HL-141797-01 and 1-UH3-HL-141797-01 to D.E.I.) and the Wyss Institute for Biologically Inspired Engineering at Harvard University (D.E.I.).

## Contributors

H.B., M.R, R.P-B. and D.E.I. conceived this study. H.B. and M.R. performed and analyzed all experiments with L.S., Y.M., J.J., R.P., C.B. and A.J. assisting with experiments and data analysis. J.D.M. and R.K.P. assisted in RNA-seq data analysis. S.R.R.H. provided assist in manuscript writing. H.B. and D.E.I. wrote the manuscript with all authors providing feedback.

## Ethical statement

All samples were de-identified and the sample number included in this study cannot reveal the identity of the study subjects. All uses of human material have been approved by the Harvard Longwood Campus Institutional Review Board.

## METHODS

### Human airway Transwell culture

Primary human lung bronchial-airway epithelial basal cells (Lonza, cat. no. CC2540; obtained from healthy donors 655308, 623950, 672447, and 626776) were expanded in 75-cm2 tissue-culture flasks using airway epithelial cell growth medium (Promocell) to 60-70% confluency. Primary diseased human lung bronchial-airway epithelial basal cells (Lonza, cat. no. 00195275; obtained from donors 18TL343179, 18TL103224, 436083, 440551, 430905) were expanded in 75-cm^2^ tissue-culture flasks using airway epithelial cell growth medium (Promocell) to 60-70% confluency. Primary human pulmonary microvascular endothelial cells (Cell Biologics) were expanded in 75-cm^2^ tissue-culture flasks using human endothelial cell growth medium (Cell Biologics) to 70-80% confluency. To create human airway transwells, 6.5mm transwells with 0.4μm pores (Sigma-Aldrich) were coated with collagen type IV from human placenta (0.5mg ml^-1^ in water; Sigma-Aldrich) at room temperature overnight. The solution was aspirated from the transwell, which was then used for seeding cells. Lung bronchial-airway epithelial basal cells (1.0 × 10^5^ cells ml^-1^) were first seeded in the apical chamber of transwell in airway epithelial growth medium (Promocell) and incubated at 37 °C with 5% CO2 until 100% confluent. The adherent cells were maintained using airway epithelial growth medium (Promocell). Once confluent, the apical medium was removed, and the basal chamber received PneumaCult-ALI medium (StemCell) to establish ALI. The airway epithelial cells were cultured for 3-4 additional weeks at 37oC with 5% CO_2_, and the apical surface of the epithelium was washed with PBS once a week to remove mucus and cellular debris. Prior to infection experiment, human pulmonary microvascular endothelial cells (1.5 × 10^6^ cells ml^-1^) were seeded on the basal side of the membrane and incubated for 2 hours at 37°C with 5% CO2. The transwells were then returned to ALI using PneumaCult-ALI media (StemCell) supplemented with 0.1% VEGF, 0.01% EGF, and 1mM CaCl_2_.

### Human Airway Chip culture

The human Airway Chip culture and infection with influenza virus was recently described (*4*). Briefly, chips were activated with ER1 solution (Emulate Inc.), washed with ER2 solution (Emulate Inc.), and then placed under a UV light (Nailstar, NS-01-US) for 20 minutes. Following that, the channels were successively washed with ER2 solution and DPBS(-/-). The porous membranes were coated on both sides with collagen type IV from human fibroblasts (0.5 mg/mL; Sigma-Aldrich) at room temperature. After aspirating the fluid, cells were seeded. Primary HLAECs (Lonza, USA) were cultivated as previously described using healthy donors (623950), COPD donors (436083 and 370751), and primary HPMVECs (Cell Biologics, USA). (*4*).

### Infection of human airway Transwell with influenza virus

Prior to infection, human airway transwells were equilibrated for 24 hours in reduced hydrocortisone (10nmol/L) differentiation media (StemCell). The airway epithelial cells were infected with influenza virus (MOI=0.1) in 50ul PBS for 1.5 hours at 37 °C with 5% CO_2_. After the incubation period, the apical solution was aspirated and the transwells were returned to 37 °C with 5% CO_2_. The apical surface of the epithelium was washed with PBS 4 hours and 18 hours post-infection and collected for analysis. The basal supernatant was collected 18 hours post-infection and analyzed for cytokines and chemokines.

### Poly(I:C) induction of human airway Transwell

Prior to induction, human airway Transwell were equilibrated for 24 hours in reduced hydrocortisone (10nmol/L) differentiation media. The apical surface of the epithelium was washed with PBS to remove mucus and cellular debris. A high molecular weight poly(I:C) stock was prepared per manufacturers specifications (Invivogen). The airway epithelial cells were then incubated apically with poly(I:C) (10ug in 50ul PBS) for 1.5 hours at 37 °C with 5% CO_2_. After the incubation period, the apical solution was aspirated and the Transwell were returned to 37 °C with 5% CO_2_. The apical surface of the epithelium was washed with PBS 4 hours and 18 hours post-infection and collected for analysis. The basal supernatant was collected 4 hours and 18 hours post-infection and analyzed for cytokines and chemokines.

### Plaque assay

Plaque assays were used to determine virus titers. Confluent MDCK cell monolayers in a 12-well plate were washed with PBS, inoculated for 1 hour at 37°C with 1 mL of 10-fold serial dilutions of samples containing influenza virus, and then overlaid with 1 mL of DMEM (Gibco) supplemented with 1.5% low melting point agarose (Sigma-Aldrich) and 2 µg/mL TPCK-treated trypsin (Sigma-Aldrich). 2-4 days after incubation at 37°C with 5% CO2, cells were fixed with 4% paraformaldehyde (Alfa Aesar) and stained with crystal violet (Sigma-Aldrich) to visualize the plaques; virus titers were measured in plaque-forming units per milliliter (PFU/mL).

### Analysis of cytokines and chemokines

The basal supernatants were collected and analyzed for a panel of cytokines and chemokines– IP-10, IL-6, IL-8, RANTES, MCP-1, TRAIL, and IL-37– using custom ProcartaPlex assay kits (Life Technologies). The analyte concentrations were determined using a Luminex 100/200 Flexmap3D instrument coupled with GraphPad Prism software.

### qPCR

Viral RNA was isolated using the QIAamp Viral RNA Kits (Qiagen). After determining RNA concentrations using a NanoDrop 2000 (Thermo Fisher), 100 ng of total RNA was used for cDNA synthesis. Reverse transcription was conducted using the Omniscript RT Kit (Qiagen). Quantitative real-time PCR was performed using the SsoAdvanced-Universal SYBR Green Supermix (Biorad). The specificity of primers was confirmed by melting curve analysis and gel electrophoresis. qPCR was performed on a CFX Connect Real Time PCR Detection System (Biorad). The primers for viral nuclear protein (NP) are: TCAAGTGAGAGAGAGCCGGA (forward) and TCAAAGTCGTACCCACTGGC (reverse). Relative viral titers were determined using the delta (Ct) method by comparing with cycle numbers of the infected control for each donor.

### Viral stocks

Influenza virus strains used in this study include A/WSN/33 (H1N1) and A/Hong Kong/8/68/ (H3N2). All viruses were obtained from the Centers for Disease Control and Prevention (CDC) or kindly shared by Drs. P. Palese, R.A.M. Fouchier, and A. Carcia-Sastre. Influenza virus strains were expanded in MDCK.2 cell as previously described (*30*).

### Drug study

At 24 hours before infection, 1 µM oseltamivir acid (Toronto Research chemicals), 1 µM or 10 µM nafamostat (Abcam) were added to transwell basal medium with a final concentration of DMSO at 0.5%. The basal medium, which contains the drug, was renewed at the time of infection. Viral load or immunofluorescence staining was performed 48 hours post infection.

### Immunostaining and confocal microscopy

Cells were rinsed with PBS(-/-), fixed with 4% paraformaldehyde (Alfa Aesar) for 20 min at room temperature, permeabilized with 0.1% Triton X-100 (Sigma-Aldrich) in PBS (PBSX) for 10 min, blocked with 5% goat serum (Life Technologies, #50062Z) in PBSX for 1 h at room temperature, and incubated with antibody diluted in blocking buffer (5% goat serum in PBSX) overnight at 4 °C, followed by incubation with fluorescent-conjugated secondary antibody for 1 h at room temperature; nuclei were stained with DAPI (Invitrogen) after secondary antibody staining. The following antibodies were used: ZO-1 Monoclonal Antibody (ZO1-1A12), Alexa Fluor 555, (Invitrogen, #MA3-39100-A555); MUC5AC Monoclonal Antibody (9-13M1), Thermo Fisher, MA1-35708. Fluorescence imaging was conducted using a confocal laser-scanning microscope (SP5 X MP DMI-6000, Germany) and image processing was done using the ImageJ software.

### RNA-seq and bioinformatic analysis

RNA-seq was performed at the Molecular Biology Core Facilities at Dana-Farber Cancer Institute using a standard RNA-seq package that includes polyA selection and sequencing on an Illumina HiSeq with 150-bp pair-ended reads. Trimmomatic v.0.36 was used to trim sequence reads to remove potential adaptor sequences and low-quality nucleotides. Using the STAR aligner v.2.5.2b, the trimmed reads were mapped to the Homo sapiens GRCh38 reference genome. We estimated unique gene hit counts using feature Counts from the Subread program v.1.5.2 and then performed differential expression analysis using DESeq2. Gene Ontology analysis was performed using DAVID (*31*) and the Enrichplot R package. Volcano plots were created using the R package EnhancedVolcano. The ggplot2 R tool was used to generate heatmaps and scatter plots. Raw data has been deposited to Gene Expression Omnibus (GEO) with the accession number GSE186908.

### Statistics

Data are displayed as mean values ± standard error of mean (SEM) unless noted otherwise in the figure legends. Graphing and statistical comparison of the data were performed using Prism 9 (GraphPad Software) or R (version 4.2) using the ggplot2 package (version 3.3.5). Unless otherwise noted, two-group comparisons were assessed using the two-tailed Student’s t test or the Mann–Whitney U test (nonparametric); comparison of three or more groups were analyzed by one-way ANOVA with Bonferroni multiple comparisons or the Wilcoxon signed-rank test (nonparametric). P values less than 0.05 were considered to be statistically significant (*, P < 0.05; **, P < 0.01; ***, P < 0.001; n.s., not significant).

## Supplementary Files

Supplemental Material available at: https://figshare.com/s/d2a7ea8ade84e2764837

Supplementary Table 1 available at: https://figshare.com/s/0f2ec5bc8113080c3150

---

**Fig. S1.**
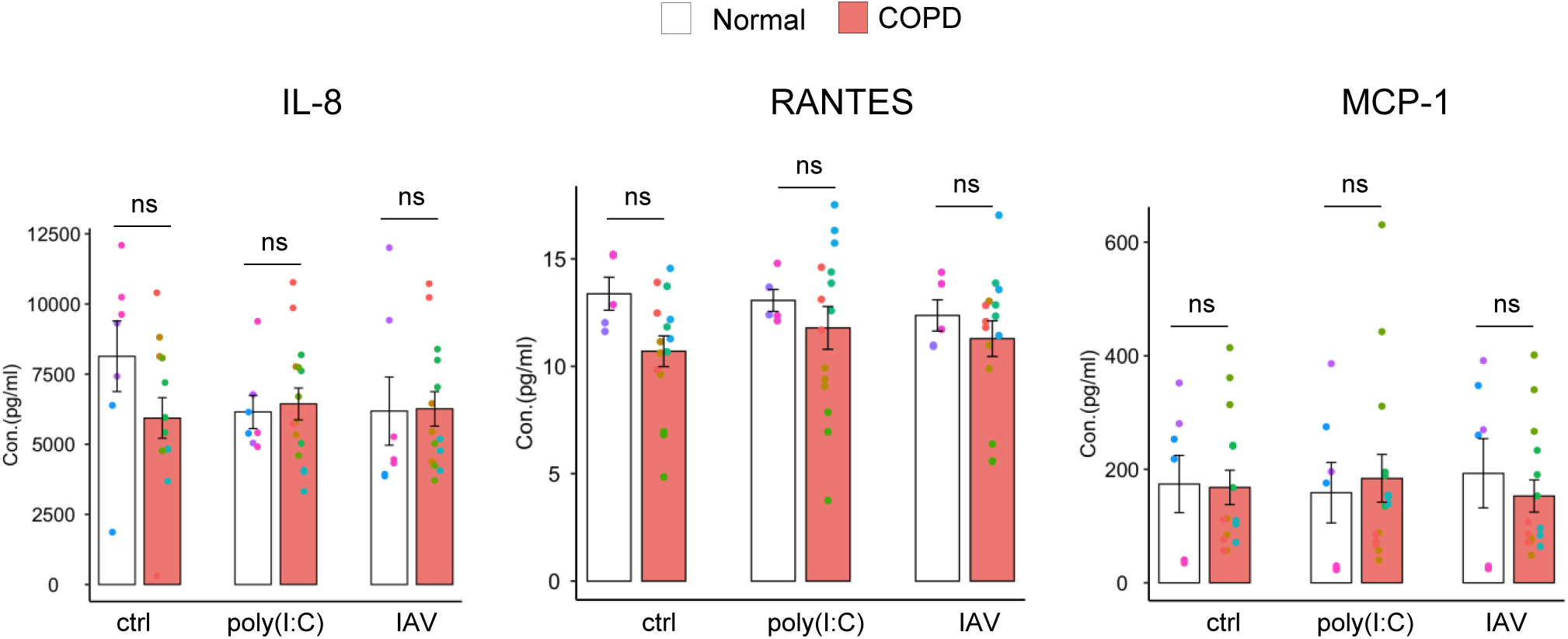
Cytokine production measured in the basal medium. ANOVA and Šidák multiple comparisons test. ns, not significant.

**Fig. S2.**
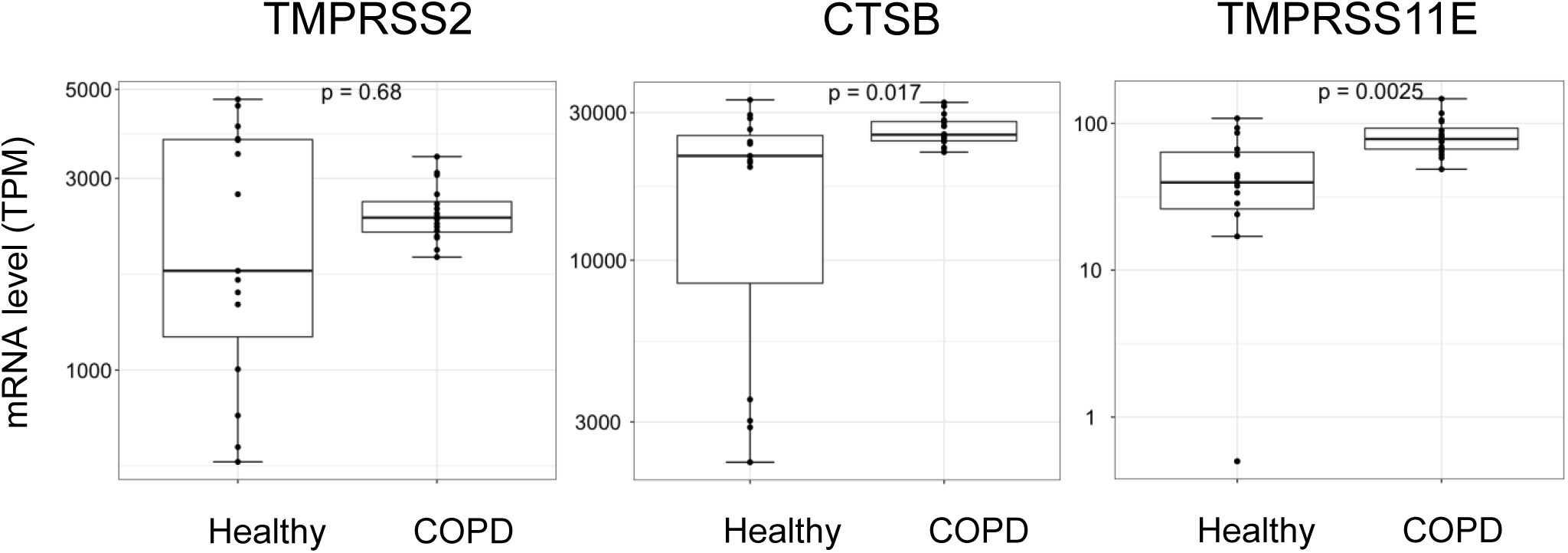
mRNA levels of influenza-activating host protease TMPRSS2, CTSB and TMPRSS11E. Mann-Whitney test.

**Fig. S3.**
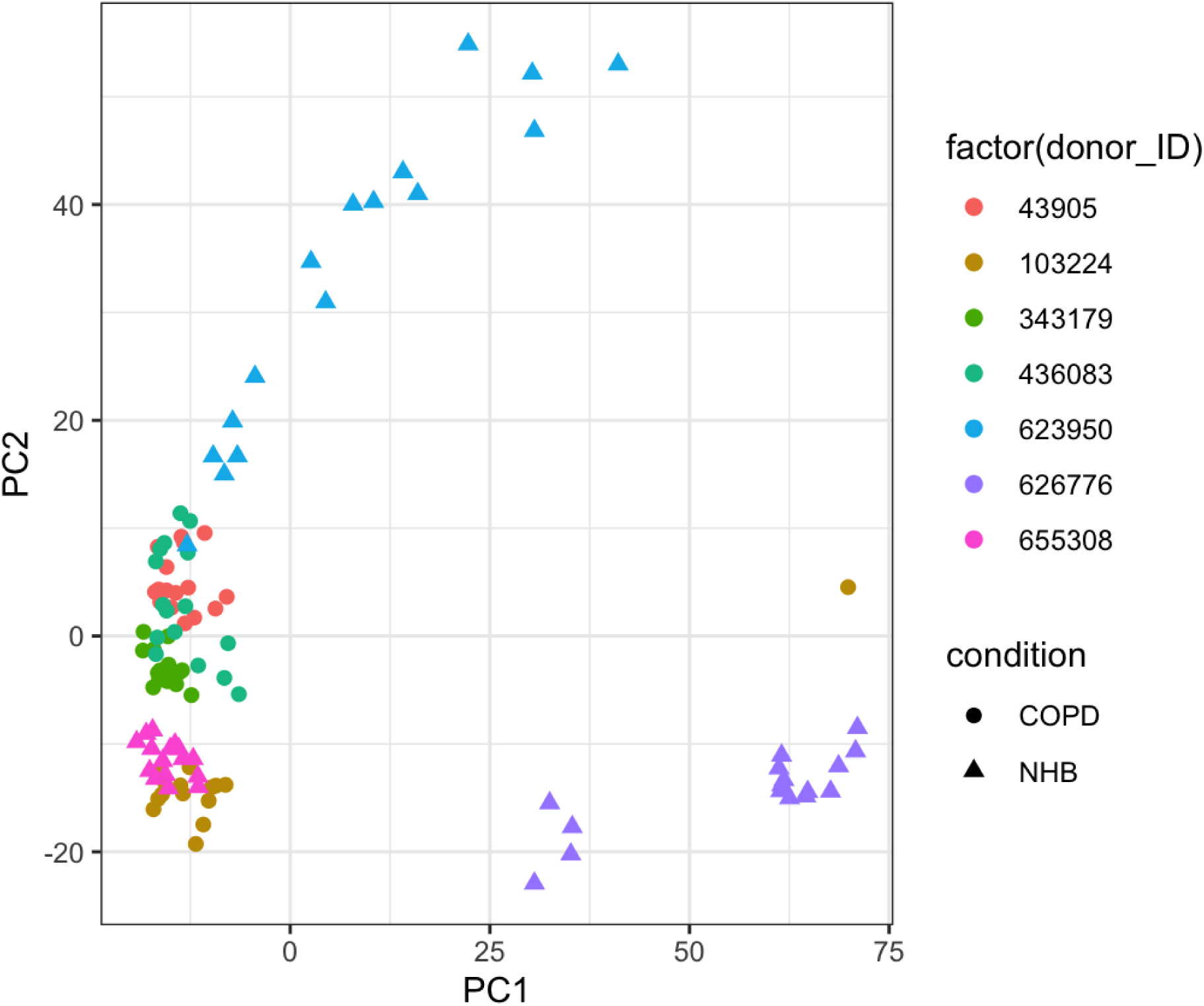
Principal component analysis (PCA) showing sample variations among healthy and COPD donor epithelium.

**Table S1.**
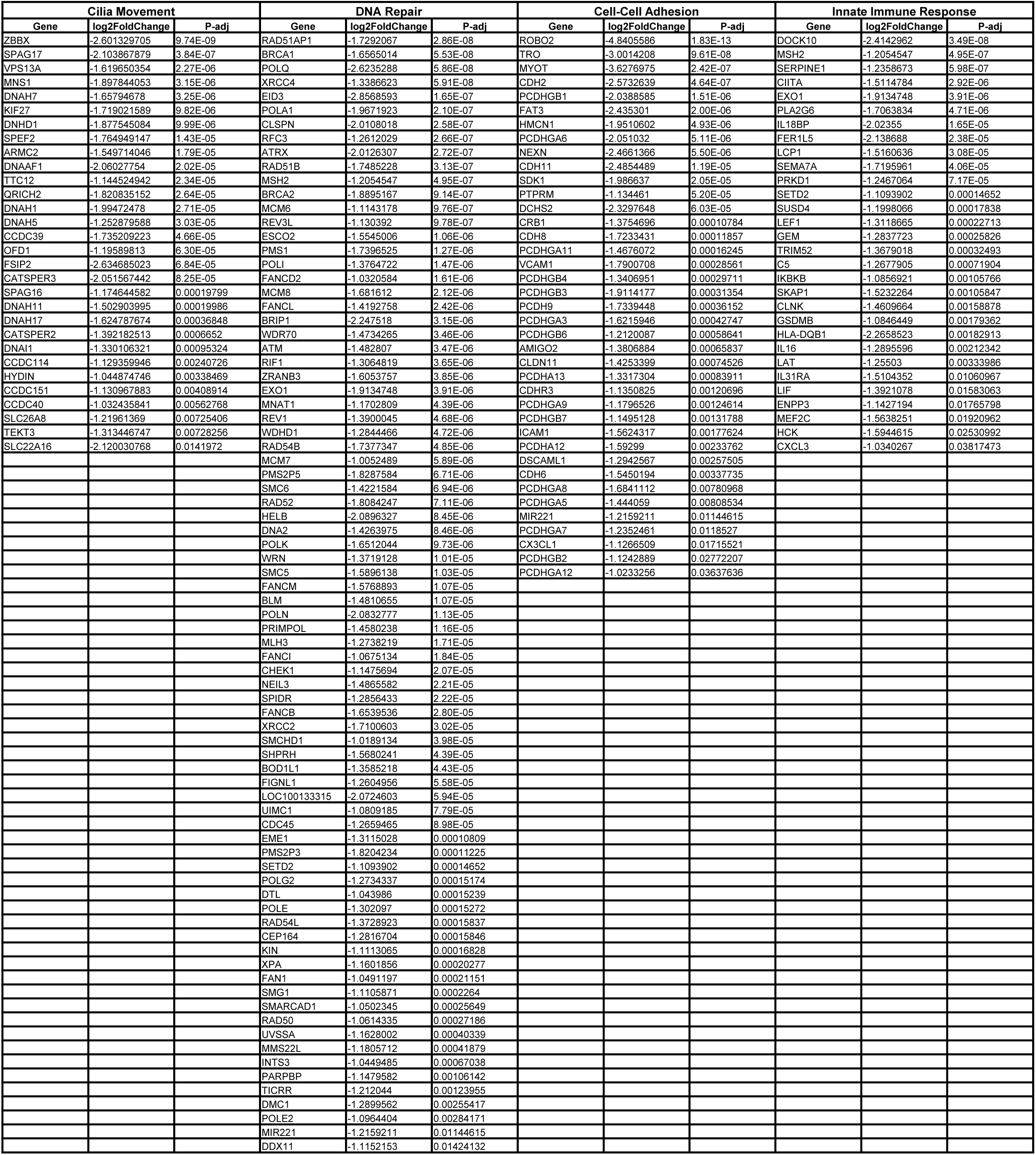
Differentially expressed genes in COPD epithelium.

